# Deep Learning for Perfusion Cerebral Blood Flow (CBF) and Volume (CBV) Predictions and Diagnostics

**DOI:** 10.1101/2023.04.20.23288688

**Authors:** Salmonn Talebi, Siyu Gai, Aaron Sossin, Vivian Zhu, Elizabeth Tong, Mohammad R. K. Mofrad

## Abstract

Dynamic susceptibility contrast magnetic resonance perfusion (DSC-MRP) is a non-invasive imaging technique for hemodynamic measurements. Various perfusion parameters, such as cerebral blood volume (CBV) and cerebral blood flow (CBF), can be derived from DSC-MRP, hence this non-invasive imaging protocol is widely used clinically for the diagnosis and assessment of intracranial pathologies, including tumor classification, stroke assessment, seizure detection, etc. Currently, most institutions use commercially available software to compute the perfusion parametric maps. Conventionally, the parametric maps are derived by mathematical equations which require the selection of vascular input waveforms. However, these conventional methods often have limitations, such as being time-consuming and sensitive to user input, which can lead to inconsistent results; this highlights the need for a more robust and efficient approach like deep learning. Using relative cerebral blood volume (rCBV) and relative cerebral blood flow (rCBF) perfusion maps generated by an FDA-approved commercial software, we trained a multi-step deep learning (DL) model. The model used each 4D MRP dataset as input, and combined the temporal features extracted from each voxel with spatial information of the brain to predict voxel-wise perfusion parameters. DL-generated rCBV and rCBF maps were evaluated qualitatively and quantitatively. An auxiliary (control) model, with similar architecture, but trained with truncated datasets that had fewer time points, was designed to explore the contribution of temporal features. Our model is based on a multistage encoder-decoder architecture that leverages a 1D convolutional neural network (CNN) as the first encoder to capture temporal information, followed by a 2D U-Net encoder-decoder network to process spatial features. This combination of encoders allows our model to effectively integrate time-varying and spatial data, generating accurate and comprehensive CBV/CBF predictions for the entire brain volume. Our model demonstrates comparable results to that of FDA-approved commercial software.

## Introduction

Going beyond anatomic imaging, magnetic resonance perfusion (MRP) imaging can provide hemodynamic information of the brain. MRP is commonly used in a wide range of clinical applications, including the assessment of stroke, tumor grading, differentiating tumor mimics, treatment decisions, etc. [1]. Dynamic susceptibility contrast (DSC) is a common non-invasive MRP technique. In DSC, dynamic T2*-weighted images are rapidly acquired after injecting a small gadolinium bolus, to capture the concentration of gadolinium within the brain every 1 to 2s. The signal intensity-versus-time curve at each voxel is inversely related to the concentration of gadolinium, which in turn, is proportional to the amount of blood in that voxel.

Various perfusion parameters, such as relative cerebral blood volume (rCBV) and cerebral blood flow (rCBF) can be derived from DSC images. Commercial software packages are often utilized to compute the perfusion maps. The underlying algorithms of these software tools are often proprietary, but most of them are based on a 2-compartment perfusion model. Within the premise of this model, the generation of the perfusion parameters require 2 vascular references – arterial input function (AIF) and venous output function (VOF). In general, cerebral perfusion parameters are dependent with a number of factors including cardiac output, caliber of the vasculature, bolus size, and injection rate. This dependence is minimized by deconvolving the arterial input function (AIF) from the tissue intensity-time waveforms. The derived perfusion parametric maps may differ according to the waveform of the AIF. The lack of consensus in the optimal choice of the AIF further impairs the reliability of the parametric maps. Similarly, the VOF is used to compensate for partial volume effect in the AIF. Again, the choice of VOF will introduce variability in the parametric maps. It is beneficial to find an efficient method to generate perfusion parametric maps that are reproducible and reliable.

Deep learning (DL) is a rapidly emerging technique which has been increasingly applied to various fields in medicine such as cardiology [2], radiology [3-5], ophthalmology [6-7], dermatology [8-9], and pathology [10-11]. Deep learning algorithms may learn from examples and respond to new inputs based on their prior training [12]. Deep learning may provide a means to generate perfusion parametric maps that are reproducible and reliable and can facilitate the assessment of stroke, tumor grading, differentiating tumor mimics, treatment decisions etc. The objective of the present study is to use deep learning neural networks to produce perfusion parametric maps with the explicit choice of an arterial input function (AIF). This will eliminate the influence of AIF and produce perfusion parameters that reflect the (patho)physiological hemodynamics more reliably. Using perfusion maps generated by an FDA approved software as ground truth, we trained DL models to predict rCBV and rCBF maps in patients with conditions associated with abnormal perfusion such as epilepsy [13], tumor [14], infectious or inflammatory, etc.

The simultaneous spatial and time-resolved nature of DSC complicated this task of predicting parametric maps. Conventional methods, such as using a convolutional neural network (CNN), on volumetric data can be memory intensive and challenging for the model to learn. In this study, we propose a two-encoder approach that first encodes the one-dimensional waveforms and then encodes the spatial information using a convolutional network with the U-Net architecture. This method significantly speeds up training and reduces memory requirements.

## Materials and Methods

This retrospective study was conducted with the approval of the Stanford Institutional Review Board (IRB) and under a waiver of informed consent. The study was approved for collaboration between Stanford University and the University of California, Berkeley. We retrospectively collected patients who underwent perfusion MRI brain imaging for clinical work-up between and found to have abnormal elevated rCBV or rCBF. Using keywords ‘hyperperfusion’, ‘hypoperfusion’, and ‘abnormal perfusion’ to search the radiologic imaging archive, resulted in 158 patients. An additional 22 patients with no perfusion abnormality were collected using keywords ‘no perfusion abnormality’ or ‘normal perfusion’. Out of the 180 patients, 21 studies were excluded due to motion artifacts. The underlying diagnoses of the 159 patients (74 females and 85 males, mean age 69 ± 11 years, range 58–80 years) are summarized in Table 1.

**Table 1.**
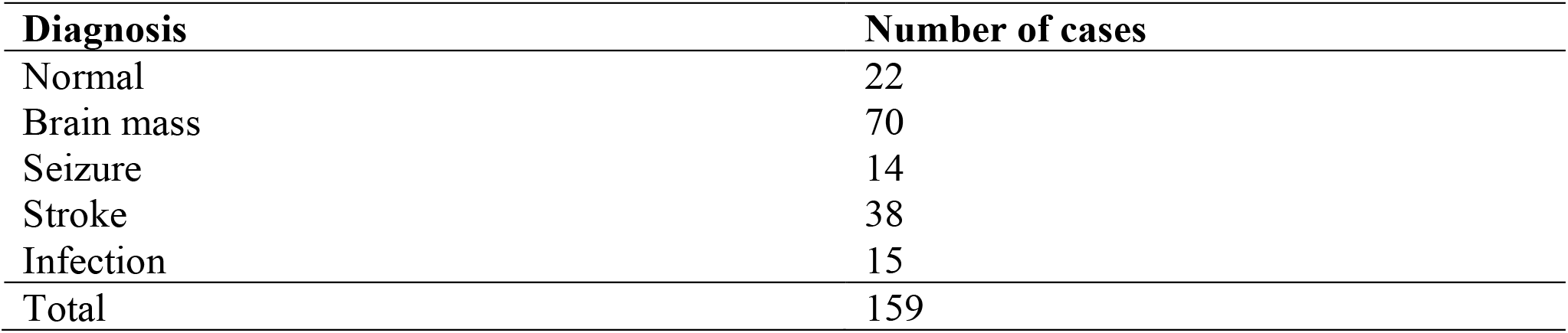
Summary of the underlying clinical diagnoses of the subjects used in this study.

### MR imaging acquisition protocol

Brain MRI was performed on a 3.0T GE MRI scanner using an 8 channel GE HR brain coil (GE Healthcare, Milwaukee, Wisconsin). DSC-MRP was performed using a dynamic susceptibility contrast technique following the intravenous administration of Multihance (Bracco, Milan, Italy) into an antecubital vein at a rate of 4.0 ml/s using a power injector. DSC parameters were: TR=1800 ms, TE=35 ms; flip angle 80°, spatial resolution=128 × 128, and slice thickness of 5 mm. A total of 60 whole-brain volumes were acquired over approximately 2-3 minutes. Relative cerebral blood flow (rCBF) and relative cerebral blood volume (rCBV) were generated by the RAPID software (iSchemaView, Menlo Park, CA) [15].

### Data Preprocessing

The MRP datasets were divided in 131 patients for training and 28 for testing. There were 60 time-samples for each subject, with one whole brain-volume for each time-time-sample. The number of slices needed to cover the entire brain of each subject varied from 18 to 22 slices. Both training and validation datasets consist of a similar mix of patients within each of the diagnostic categories.

Raw perfusion scans of patients were saved as DICOM files and were imported as inputs to our models. rCBV and rCBF parametric files produced by the commercial software were imported the ground truth outputs. Pixel intensities range from 0 to 17199. Higher pixel intensities indicate higher rCBV or rCBF. A data pipeline was built to import and preprocess the data before inputting it into the model. DSC images went through skull stripping and co-registration. The images and the perfusion maps then went through clipping, normalization, and standardization steps. Inputs to the model were separated slice-wise for each subject and were aggregated afterwards to generate the final prediction.

### Model Architecture

We designed a multistage encoder architecture followed by a decoder to process the MR Perfusion scans (Figure 1). A 1-D convolutional neural network is used as the first encoder which integrates information over the time dimension. Extracted features from the CNN encoder block are then reshaped and passed to a 2D encoder-decoder U-Net network. The U-net architecture performed multiple down-sampling steps on the input to extract important features. Multiple up-sampling steps were then performed. The up-sampled features were then concatenated with the outputs of the down-sampling layers. The output of the U-net further was passed through two additional 2D-CNN layers to extract parametric maps. Finally, the predicted slices were then recombined into a final volumetric output containing the CBV/CBF predictions for the patient’s entire brain volume. This full dataset, comprising of 60 time-points, was utilized in the training of this model (*model_full*). To explore the importance of temporal features, we adapted from the *model_full* and trained another model (*model_truncated*) with only the first 40 time-points.

**Figure 1:**
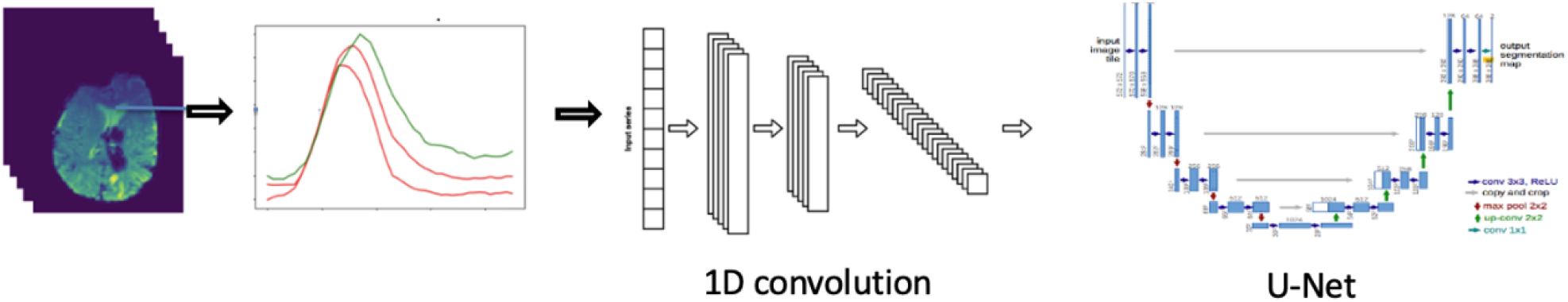
We designed a multistage encoder architecture followed by a decoder to process the MRP datasets. The model captures temporal features followed by a modified U-net to generate the perfusion maps.

### Model training and evaluation

The training patient data were randomly divided into two sets: training (70%) and validation (30%). A separate test set containing 28 patients was used as a final evaluation for our best performing model. The validation set was used for performing a hyperparameter grid search, specifically tuning the learning rate from a range of 1×10^−4^ to 1×10^−6^. The models converged after training for 200 epochs. After fine-tuning the model with the validation set, we evaluated the best performing model on our test set. The models were trained using a single A6000 GPU.

The predicted rCBV and rCBF maps were qualitatively assessed by two neuroradiologists for quality assurance. Each reader evaluated the rCBF and rCBV maps in a randomized order and evaluated them for global image quality and for adequate differentiation between white and gray matter. For quantitative assessment, the mean absolute error (MAE) and root mean square error (RMSE) were calculated using the maps generated by the commercial software as the ground truth.

## Results

The predicted rCBV and rCBF maps were displayed in color and evaluated by two neuroradiologists. Representative results from one test subject are illustrated in Figure 2. Qualitatively, all the predicted rCBV and rCBF maps from the test set were deemed diagnostic. There were small areas of under-estimated rCBV and rCBF values in the maps predicted by *model_full* and *model_truncated*. There were more under-estimated pixels in the maps predicted by *model_truncated*.

**Figure 2.**
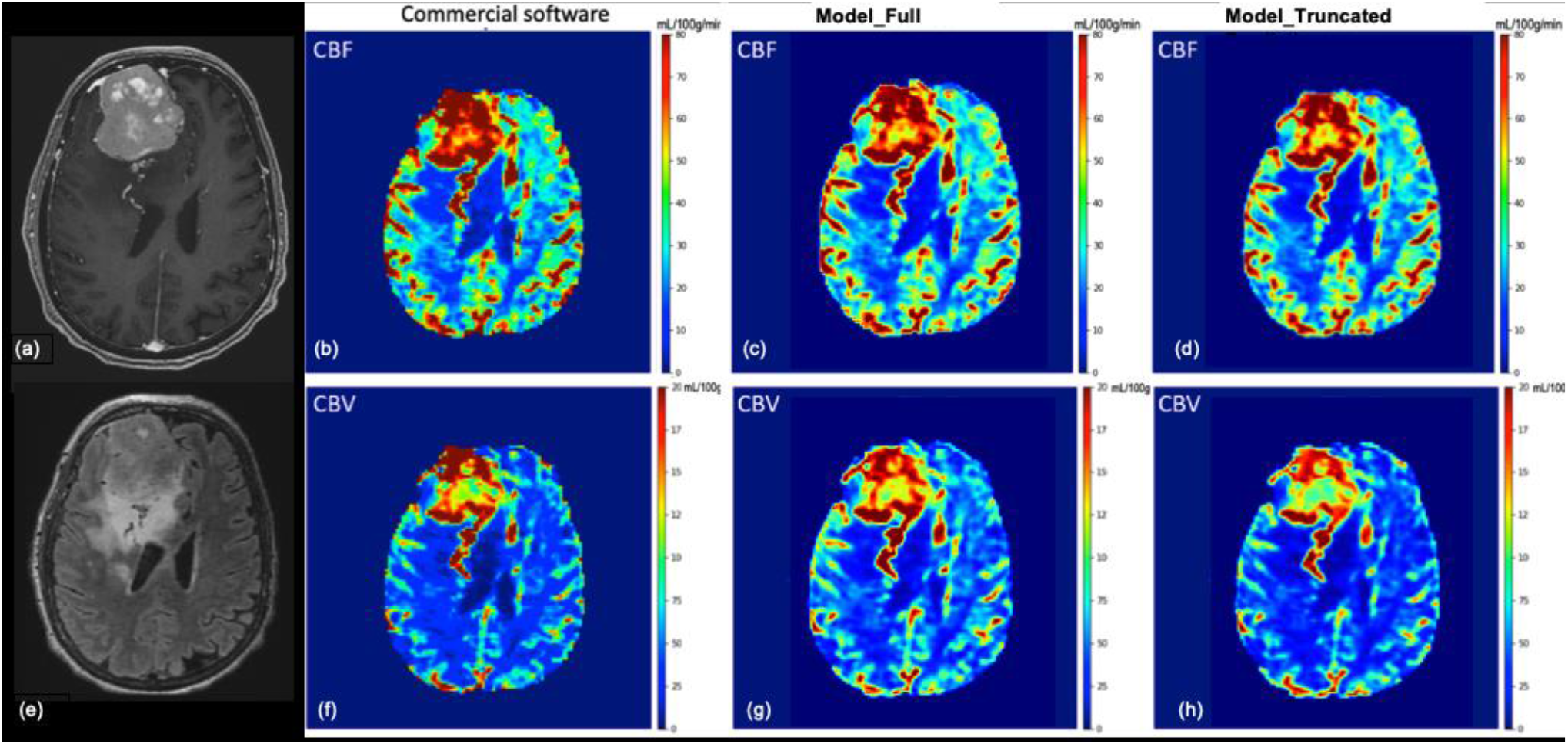
Female in their 80’s with (a) right frontal WHO grade 2 meningioma, measures approximately 5.6 × 6.9 × 4.8 cm with heterogeneous avid enhancement, and surrounding edema. Mass compressed right lateral ventricle and caused midline shift to the left. A few engorged medullary veins developed along the posterior margin of the mass, draining into cortical and medullary veins. (e) There was T2/FLAIR hyperintense edema around the mass. (b) rCBF and (f) rCBV maps calculated by a FDA-approved software. There was markedly elevated rCBF within the mass, there was elevated rCBV within the mass especially along the anterior and posterior aspects. (c) rCBF and (g) rCBV maps predicted by the model_full, which was trained with 60 time-points. The model_full predicted elevated rCBF within the mass, and elevated rCBV along the anterior and posterior aspects. There were small areas where the predicted rCBF and rCBV were under-estimated. (d) rCBF and (h) rCBV maps predicted by the model_truncated, which was trained with 40 time-points slightly under-estimated the rCBF and rCBF values in a few small areas. (b, f) Note, there was also elevated rCBF and rCBV within the engorged medullary draining veins, which were consistently predicted by both model_full and model_truncated. Overall, the predicted rCBV and rCBF maps were diagnostic and useful.

**Figure 3.**
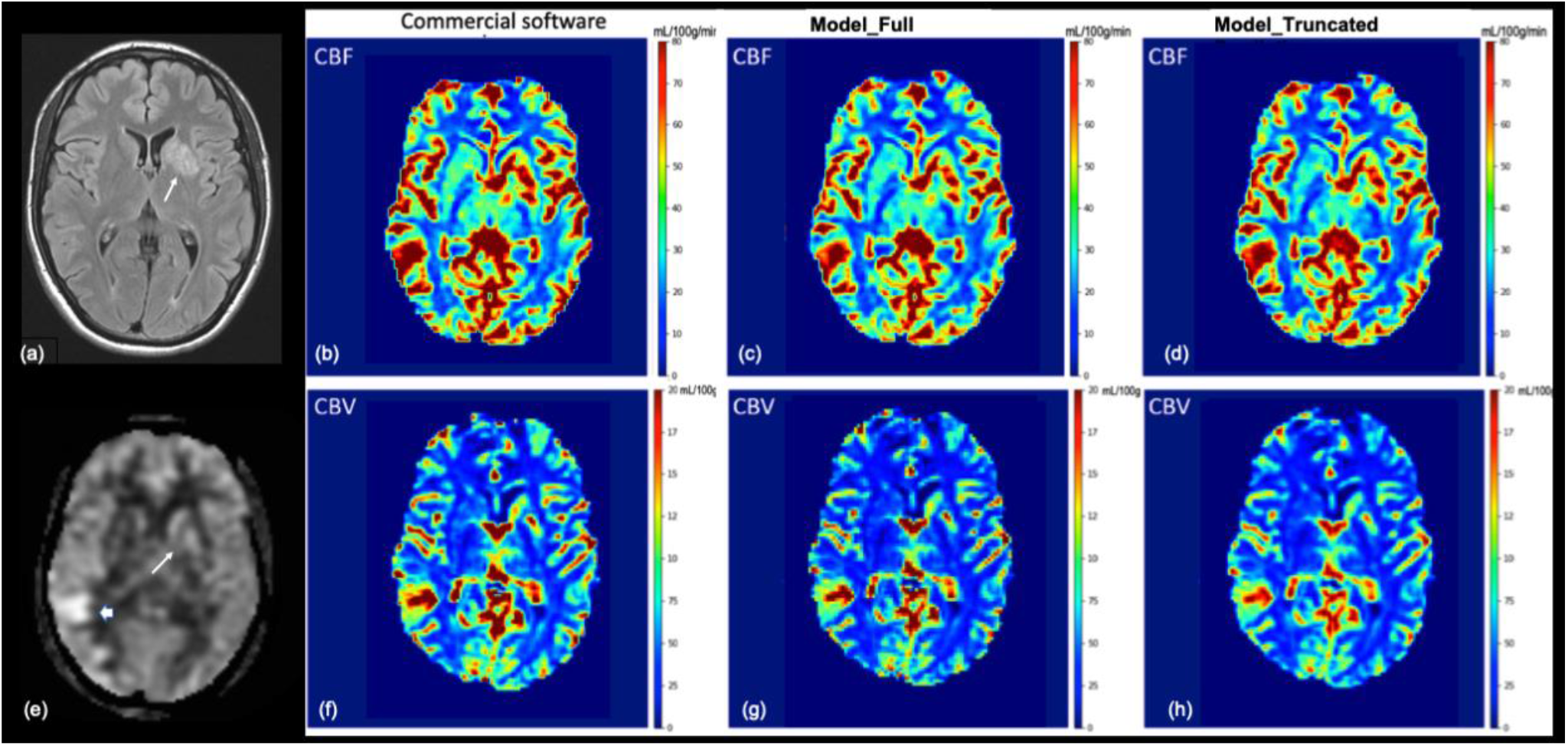
Male in their 20’s presented with seizure with focal neurologic deficits, diagnosed with autoimmune encephalitis. (a) There was a FLAIR T2 hyperintense lesion in left basal ganglia (white arrow). (e) There was associated hyperperfusion in left basal ganglia (white arrow) on ASL (arterial spin labeling). There was also a small area of confluent hyperperfusion in right temporal lobe (thick white arrow) likely related to seizures. (b) rCBF and (f) rCBV maps calculated by a FDA-approved software. There was confluent elevated rCBF within the left basal ganglia and right temporal lobe. There were small areas of elevated rCBV within the left basal ganglia and right temporal lobe. (c) rCBF and (g) rCBV maps predicted by the model_full, which was trained with 60 time-points. The model_full predicted elevated rCBF within the left basal ganglia and right temporal lobe. (d) rCBF and (h) rCBV maps predicted by the model_truncated, under-estimated the rCBF and rCBF values in a few small areas.

The performance metrics of the *model_full* (trained on 60 time-points) and *model_truncated* (trained on 40 time-points) from the test set were tabulated in Table 2. Using the whole dataset, the mean MAE and RMSE for the rCBF maps were 1.91mL/100g/min and 7.77mL/100g/min respectively; the mean MAE and RMSE for the rCBV maps were 0.59 mL/100g and 2.63 mL/100g, respectively. The performance took a slight hit when the model was trained on fewer time-points. The *model_truncated* scored a slightly lower mean MAE and RMSE for the rCBF maps of 3.11 mL/100g/min and 10.88 mL/100g/min respectively; the mean MAE and RMSE for the rCBV maps were 0.85 mL/100g and 3.53 mL/100g, respectively.

**Table 2.** Performance of the *model_full* (trained on 60 time-points) and *model_truncated* (trained on 40 time-points) from the test set.

|  | Model_Full |  | Model_Truncated |  |
| --- | --- | --- | --- | --- |
|  | MAE Loss<br>(mL/100g/min) | RMSE<br>(mL/100g/min) | MAE Loss<br>(mL/100g/min) | RMSE<br>(mL/100g/min) |
| rCBF | 1.91 | 7.77 | 3.11 | 10.88 |
| rCBV | 0.59 | 2.63 | 0.85 | 3.53 |

## Discussion

Dynamic susceptibility contrast magnetic resonance perfusion (DSC-MRP) have a wide range of clinical applications, including the classification of tumors, identification of stroke regions, and characterization of other diseases such as cancer tumors etc. rCBV and rCBF maps are especially useful in hyper-perfused states, such as epilepsy, tumor, infectious or inflammatory, etc.

Deconvolution is most frequently used MRP postprocessing method, it can be either based on singular value decomposition or Fourier transform [16]. Both post-processing techniques are sensitive to artifacts and noise, and may introduce noise in the estimated output parametric maps. Deconvolution methods require the selection of two vascular waveforms - the arterial input function (AIF) and the venous output function.

There is no consensus in the optimal choice of AIF and VOF. The time-intensity waveform of the AIF can differ based on the artery selected, as a result, the parametric maps may vary according to the selection of AIF. Similarly, the parametric maps may also vary according to the choice of VOF. When performed manually, the choice of AIF and VOF can be affected by noise and operator’s experience. Poor reproducibility of vessel selection will result in unreliable perfusion maps. It is, therefore, desirable to develop a method that is robust to noise and the choice of vascular functions.

In recent years, deep learning (DL) models have been shown to be a valuable technique in radiology. Besides using DL for detection and segmentation intracranial abnormalities in brain images [17-18], there are also DL models for predictions from input images [19-20]. DL methods have also been used in MRP. There are commercial MRP software that have built-in automated AIF and VOF selection tools that utilize DL [21]. Although the underlying algorithms of most the commercial software are proprietary, other groups have published their algorithm [22-24]. Using DL methods to select the AIF and VOF were more reproducible and less operator-dependent. Once AIF and VOF were selected, the perfusion parameters are calculated by conventional convolution methods. As such, DL is only limited to the selection of vascular input functions. McKinley et al. took this further and used DL to generate perfusion maps by comparing the voxel-wise intensity-time waveform with the pre-selected AIF waveform [25]. They compared several machine learning techniques such as random forests, linear regression and neural network to generate the perfusion parametric maps. Our approach does not require pre-selection of vascular input or output functions. Instead, our model implicitly incorporates the arterial input waveforms and venous output waveforms as inherent variables embedded within each 4D-dataset.

Traditional perfusion post-processing operates on a voxel level. Information from neighboring voxels may be incorporated by applying a spatial smoothing filter before the deconvolution step. In our approach, temporal features and spatial information are very purposefully and deliberately combined together to generate the perfusion maps. In our two-step encoder approach, temporal features are first extracted from each voxel. In the second stage, these temporal features are then integrated with spatial features through a U-Net neural network. To illustrate the importance of the foremost step of temporal feature extraction, we trained a model, with similar architecture, with truncated datasets. As expected, this resulted in a small hit in the performance of the model. Conventionally, cerebral blood volume is obtained by integrating the area under the deconvolved tissue concentration-time curve [26]. A ML model trained with truncated waveform will likely produce less accurate results. Similarly, cerebral blood flow, which is estimated by the slope of the deconvolved tissue concentration-versus-time curve, will also be impacted by shortened waveforms. Our findings confirm that temporal features of the intensity-time waveforms have important impact on the overall performance.

Deep learning architectures have demonstrated utility to be an effective strategy for generating parametric mapping from perfusion scans. This DL-based method may act as an affordable and dependable alternative for generating perfusion parametric maps for the assessment of acute ischemic stroke, brain tumors, and other diseases.

Our current model demonstrates comparable results to that of an FDA-approved commercial software. Quantitatively, our model gives comparable results to the ground truth maps. Qualitatively, our model generated rCBV and rCBF maps that are good enough for a radiologist to provide a clinical assessment of the patient. The model performed well in delineating the pathologies. There were small areas within the lesion where the absolute values were under-estimated. This is likely due to the small training set. Although this did not affect the diagnostic quality of the maps, it would lead to errors in quantitative measures. Further fine-tuning of the model is needed on our current model.

## LIMITATIONS

Our study has several limitations. The generalizability of the model is limited since our data came from a single center. Therefore, further studies with multicenter data will be needed to evaluate how susceptible our model is to distribution shift. Larger number of datasets may also improve the performance.

## CONCLUSION

We demonstrate that DL models can produce perfusion maps from source perfusion imaging. By bypassing the selection of vascular input or output functions, the parametric maps are highly reproducible and operator independent. Detection or segmentation models can be built upon this model to provide end-to-end automated perfusion analysis of different neurologic pathologies.

## Data Availability

The datasets utilized during this study are not publicly available due to reasonable privacy and security concerns. The data is not easily redistributable to researchers other than those engaged in the Institutional Review Board-approved research collaborations with Stanford University.

## AUTHOR CONTRIBUTIONS

ET conceived of the research study. SG and ST contributed toward the design, implementation and evaluation of deep learning models for CBV/CBF model prediction. AS, SG, ST, VZ contributed toward the data pipeline and preprocessing techniques. ET, MM, SG, ST managed the project vision and implementation along with writing of the manuscript.

## REFERENCES

1. Tong E, Sugrue L, Wintermark M. Understanding the Neurophysiology and Quantification of Brain Perfusion. Top Magn Reson Imaging TMRI. 2017;26(2):57–65. doi:10.1097/RMR.0000000000000128

2. Madani, A., Ong, J. R., Tibrewal, A. & Mofrad, M. R. K. Deep echocardiography: data efficient supervised and semi-supervised deep learning towards automated diagnosis of cardiac disease. npj Digital Med. 1, 1–11 (2018)

3. Ribli, D., Horvath, A., Unger, Z., Pollner, P. & Csabai, I. Detecting and classifying lesions in mammograms with deep learning. Sci. Rep. 8, 4165 (2018).

4. Lindsey, R. et al. Deep neural network improves fracture detection by clinicians. Proc. Natl. Acad. Sci. 115, 11591–11596 (2018).

5. Lehman, C. D. et al. Mammographic breast density assessment using deep learning: clinical implementation. Radiology 290, 52–58 (2019).

6. Gulshan, V. et al. Development and validation of a deep learning algorithm for detection of diabetic retinopathy in retinal fundus photographs. JAMA 316, 2402–2410 (2016).

7. Lee, C. S., Baughman, D. M. & Lee, A. Y. Deep learning is effective for classifying normal versus age-related macular degeneration optical coherence tomography images. Ophthalmol. Retina 1, 322–327 (2016).

8. Phillips, M. et al. Assessment of accuracy of an artificial intelligence algorithm to detect melanoma in images of skin lesions. JAMA Netw. Open 2, e1913436 (2019).

9. Han, S. S. et al. Classification of the clinical images for benign and malignant cutaneous tumors using a deep learning algorithm. J. Invest. Dermatol. 138, 1529–1538 (2018)

10. Steiner, D. F. et al. Impact of deep learning assistance on the histopathologic review of lymph nodes for metastatic breast cancer. Am J. Surg. Pathol. 42, 1636–1646 (2018)

11. Fuyong Xing, F., Hai, Su. H., Neltner, J. & Lin, Y. L. Automatic Ki-67 counting using robust cell detection and online dictionary learning. IEEE Trans. Biomed. Eng. 61, 859–870 (2014)

12. Rajkomar A, Dean J, Kohane I. Machine learning in medicine. New England Journal of Medicine. 380, 1347–1358 (2019).

13. Kim SE, Lee BI, Shin KJ, et al. Characteristics of seizure-induced signal changes on MRI in patients with first seizures. Seizure - Eur J Epilepsy. 2017;48:62–68. doi:10.1016/j.seizure.2017.04.005

14. Wong JC, Provenzale JM, Petrella JR. Perfusion MR Imaging of Brain Neoplasms. Am J Roentgenol. 2000;174(4):1147–1157. doi:10.2214/ajr.174.4.1741147

15. Straka M, Albers GW, Bammer R. Real-time diffusion-perfusion mismatch analysis in acute stroke. J Magn Reson Imaging. 2010;32(5):1024–1037. doi:10.1002/jmri.22338

16. Konstas AA, Goldmakher GV, Lee TY, Lev MH. Theoretic Basis and Technical Implementations of CT Perfusion in Acute Ischemic Stroke, Part 2: Technical Implementations. Am J Neuroradiol. 2009;30(5):885–892. doi:10.3174/ajnr.A1492

17. Liu CF, Hsu J, Xu X, et al. Deep learning-based detection and segmentation of diffusion abnormalities in acute ischemic stroke. Commun Med. 2021;1(1):1–18. doi:10.1038/s43856-021-00062-8

18. Ottesen JA, Yi D, Tong E, et al. 2.5D and 3D segmentation of brain metastases with deep learning on multinational MRI data. Front Neuroinformatics. 2023;16. Accessed March 8, 2023. https://www.frontiersin.org/articles/10.3389/fninf.2022.1056068

19. Ding Y, Sohn JH, Kawczynski MG, et al. A Deep Learning Model to Predict a Diagnosis of Alzheimer Disease by Using 18F-FDG PET of the Brain. Radiology. 2019;290(2):456–464. doi:10.1148/radiol.2018180958

20. Cole JH, Poudel RPK, Tsagkrasoulis D, et al. Predicting brain age with deep learning from raw imaging data results in a reliable and heritable biomarker. NeuroImage. 2017;163:115–124. doi:10.1016/j.neuroimage.2017.07.059

21. Lansberg M, Lee J, Christensen S, et al. Utility of Automated MRI Analysis Software (RAPID) to Select Patients for Reperfusion Therapy: A Pooled Analysis of the EPITHET and DEFUSE Studies. Stroke J Cereb Circ. 2011;42(6):1608–1614. doi:10.1161/STROKEAHA.110.609008

22. Bjørnerud A, Emblem KE. A fully automated method for quantitative cerebral hemodynamic analysis using DSC–MRI. J Cereb Blood Flow Metab Off J Int Soc Cereb Blood Flow Metab. 2010;30(5):1066–1078. doi:10.1038/jcbfm.2010.4

23. Yin J, Yang J, Guo Q. Automatic determination of the arterial input function in dynamic susceptibility contrast MRI: comparison of different reproducible clustering algorithms. Neuroradiology. 2015;57(5):535–543. doi:10.1007/s00234-015-1493-9

24. de la Rosa E, Sima DM, Menze B, Kirschke JS, Robben D. AIFNet: Automatic vascular function estimation for perfusion analysis using deep learning. Med Image Anal. 2021;74:102211. doi:10.1016/j.media.2021.102211

25. McKinley R, Hung F, Wiest R, Liebeskind DS, Scalzo F. A Machine Learning Approach to Perfusion Imaging With Dynamic Susceptibility Contrast MR. Front Neurol. 2018;9. Accessed March 8, 2023. https://www.frontiersin.org/articles/10.3389/fneur.2018.00717

26. Copen WA, Schaefer PW, Wu O. MR Perfusion Imaging in Acute Ischemic Stroke. Neuroimaging Clin N Am. 2011;21(2):259. doi:10.1016/j.nic.2011.02.007

